# Comparative Analysis of Commercially Available Typhoid Point of Care Tests: Results of a Prospective and Hybrid Retrospective Multicenter Diagnostic Accuracy Study in Kenya and Pakistan

**DOI:** 10.1101/2022.07.17.22277655

**Authors:** Jyotshna Sapkota, Rumina Hasan, Robert Onsare, Sonia Arafah, Sam Kariuki, Sadia Shakoor, Farah Qamar, Sheillah Mundalo, Frida Njeru, Rael Too, Elizabeth Ndegwa, Jason R Andrews, Sabine Dittrich

**Author notes:** Address correspondence to Jyotshna Sapkota.

## Abstract

Blood and bone marrow cultures are considered the gold standard for the diagnosis of typhoid, but those methods require infrastructure and skilled manpower that are not always available in low- and middle-income countries where typhoid is endemic. The objective of the study is to evaluate the sensitivity and specificity of nine commercially available *Salmonella* Typhi rapid diagnostic tests (RDTs) using blood culture as reference standard in a multicenter study. This was a prospective and retrospective multicenter diagnostic accuracy study conducted in two geographically distant typhoid-endemic areas (Pakistan and Kenya; NCT04801602). 9 RDTs were evaluated were including Widal test. Point estimates for sensitivity and specificity were calculated using the Wilson method. Latent class analyses were performed using R to address the imperfect gold standard. 531 serum samples were evaluated (264 blood-culture positive; 267 blood-culture negative). The sensitivity of RDTs varied widely (range 0–78.8%), with the best overall performance shown by Enterocheck WB (72.7% sensitivity: 86.5% specificity). In latent class modeling, CTK IgG was found to have the highest sensitivity (79.1%), while the highest overall accuracy was observed with Enterocheck (73.8% sensitivity and 94.5% specificity). All commercially available *Salmonella* Typhi RDTs evaluated in the study had sensitivity and specificity values that fell below the required levels to be recommended for an accurate diagnosis. There were minimal differences in RDT performances between endemic regions. These findings highlight the clear need for new, more accurate *Salmonella* Typhi tests.

## Introduction

Typhoid fever is an enteric bacterial infection caused by the bacterium *Salmonella* Typhi (*S*. Typhi), which is primarily transmitted through contaminated food or water (1). Symptoms include prolonged fever, fatigue, headache, nausea, abdominal pain, constipation or diarrhea, with severe cases leading to serious complications and even death. In 2018, it was reported that 11–20 million people worldwide contract typhoid each year, resulting in 128,000–161,000 deaths (2).

Because of the primary mode of transmission, typhoid cases are most prevalent in places with poor sanitation and a lack of safe drinking water, most commonly low- and middle-income countries (LMICs) (2). Of these, endemic regions with the highest incidence of typhoid are sub-Saharan Africa (accounting for 40% of all cases), South Asia, North Africa and the Middle East (2). While the disease can be effectively treated with antibiotics, escalating global antimicrobial resistance, including the emergence of extensively drug-resistant *S*. Typhi in Pakistan and azithromycin-resistant *S*. Typhi in multiple countries, indicates that short-to medium-term control of typhoid through vaccination may be the best strategy for reducing disease burden in endemic populations (3, 4). However, with no reliable point-of-care diagnostic, accurately measuring disease burden in order to effectively target areas where routine vaccination would provide the greatest benefit remains a challenge (4).

An accurate diagnosis of typhoid can prove difficult, as the characteristic symptoms are similar to other undifferentiated febrile illnesses such as malaria or dengue fever, and typhoid can also be mistaken for vector borne febrile illnesses such as scrub typhus (5). As such, microbiological testing is usually required to confirm a diagnosis of *S*. Typhi, with blood and bone marrow cultures currently considered the gold-standard tests (6, 7). However, blood culture testing can be expensive, has a low sensitivity, and requires infrastructure and skilled staff that are not always available in LMICs and are not adequate for rapid patient management (6, 7). Tests using bone marrow cultures may have a higher sensitivity than blood culture tests, but they are not routinely performed as obtaining bone marrow aspirates involves skilled invasive techniques (6, 7). As a result, alternative tests have been widely adopted, especially in LMICs. The most widely used rapid diagnostic test (RDT) is the Widal test, despite numerous reports of poor sensitivity and specificity, which in some cases has led to multiple misdiagnoses in disease outbreaks, treatment delays, and even deaths (6, 8–10). Limitations have also been reported for other typhoid RDTs, such as Typhidot (IDL Biotech), Tubex (Reszon Diagnostics), and Test-it(tm) Typhoid IgM (LifeAssay Diagnostics) (6). While these show improvements over the Widal test, they still only exhibit moderate sensitivity and specificity (6). However, many accuracy studies into the effectiveness of typhoid RDTs have variations in methodology and reference standards, making it difficult for healthcare providers and policymakers to make robust decisions or recommendations for the utility of commercial tests in different settings.

The primary aim of this study was to evaluate the sensitivity and specificity of various commercially available typhoid RDTs using the same protocol in different endemic settings and comparing against a single reference standard available in endemic regions. In parallel, we aimed to (i) develop a biorepository of well-characterized specimens for use in evaluating emerging diagnostic tests for typhoid and for supporting future test development, and (ii) evaluate the operational characteristics (including invalid and indeterminate testing rates) of the RDTs.

## Methods

### Study design

This was a prospective and retrospective multicenter diagnostic accuracy study to evaluate the sensitivity and specificity of commercially available typhoid RDTs, using blood culture testing as a reference standard (NCT04801602). The study was conducted in accordance with the principles of the Declaration of Helsinki, the International Conference on Harmonization of technical requirements for registration of pharmaceuticals for human use – Good Clinical Practice guidelines (ICH-GCP E6 [R2]), and all applicable local IRBs and national laws and regulations. The studies were approved by local nation IRB regulatory bodies The study was conducted across 9 study sites in two geographically distant typhoid-endemic regions (Pakistan [South Asia] and Kenya [Sub-Saharan Africa]) between October 2020 and July 2021. Prospective patients were recruited upon clinical encounters at one of the clinical study sites. Retrospective blood culture samples (that had been tested for *S*. Typhi) were collected from a previous population-based infectious disease study (SSC 2761, Population-Based Infectious Disease Surveillance (PBIDS): A Platform to measure disease burden and evaluate the impact of public health interventions in Asembo, Siaya County and Kibera, Nairobi County, Kenya) to ensure an adequate sample size for analysis. The aim of the study was to establish a PBIDS platform suitable for tracking causes and burden of common infectious diseases such as typhoid fever over time in both urban and rural settings.

### Patients

#### Prospective enrolment

Patients eligible for prospective inclusion in the study were 2–65 years of age (8–65 years in Kenya), presenting with fever or an axillary temperature of >37.5°C (or a history of 3 or more days of fever in the week prior to enrolment), a clinical suspicion of enteric fever, and a body weight >8 kg, who had been admitted to hospital within the past 12 hours or had presented to the outpatient department or emergency department of the participating study site.

Patients were excluded from the study if they were unable or unwilling to provide the required blood samples for analysis, were receiving anticoagulant medications, did not provide informed consent, or had a known non-infectious or non-typhoid-related cause of fever.

#### Retrospective enrolment

Retrospective serum samples (from study SSC 2761) included in the study were from febrile patients (<7 days), if sample volume was >0.5 mL, had been stored at or below −20°C, were non hemolytic, and had been through less than three freeze-thaw cycles. From the parent study (SSC 2761), all residents of the two study sites, Asembo and Kibera, Kenya were included for household surveillance upon consent from the household head. For the health facility (clinic) surveillance at St. Elizabeth Lwak Mission Hospital in Asembo and Tabitha Medical Clinic in Kibera, patients who were residents of the respective PBIDS areas were included in the study if they visited the facility and fulfilled the case definition of the target infectious diseases (11).

### Investigational products and study procedures

Upon enrolment in the study, blood samples were obtained from each patient by trained phlebotomists using venipuncture, transported to a central laboratory within 24 hours (Aga Khan University in Pakistan; Centre for Microbiology Research [CMR] at Kenyan Medical Research Institute [KEMRI] in Kenya) and blood culture was carried out. *S*. Typhi RDTs were carried out once blood culture results were available, samples were stored at −20°C or below until analyzed. Readers of RDTs were blinded to the results of the blood culture tests and *vice versa*.

Following testing, any remaining serum samples were aliquoted and stored at −80°C for future evaluation of diagnostic tests (via the https://www.finddx.org/biobank-services/specimen-bank/mrf/)

#### Rapid diagnostic tests

For RDTs, 4 mL and 3 mL of blood was drawn from adult patients and children, respectively. Following transportation to the central laboratories, serum was separated and RDTs performed according to manufacturer’s instructions by trained laboratory personnel.

Nine typhoid RDTs were selected for use in the study, based on their quality (must be CE-marked), published data, international availability, required sample volume, cost, and turnaround time. Of the nine tests, one measured the presence of *S*. Typhi and/or Paratyphi antigens (Diaquick S. typhi/paratyphi Ag cassette [Dialab]; only used in Pakistan), four measured immunoglobulin (Ig) G and IgM anti-*S*. Typhi antibodies (SD Bioline *S*. Typhi IgG/IgM Fast [Abbott]; Typhidot Rapid IgG/IgM combo test [Reszon Diagnostics International]; Typhoid IgG/IgM Combo Rapid Test CE [CTK Biotech]; Typhoid IgG/IgM Rapid Test Cassette [Spectrum Diagnostics]), and three measured just the IgM antibodies (TUBEX TF [IDL Biotech]; Enterocheck WB [Tulip diagnostics]; Test-it™ Typhoid IgM [Life Assay]). The Widal test, which measures the presence of ‘O’ and ‘H’ anti-*S*. Typhi and *S*. Paratyphi antibodies, was also included due to its widespread use in endemic regions.

#### Blood culture tests

For blood culture tests, 10 mL and 3.5 mL of blood was drawn from adult patients and children, respectively. Following transportation to the central laboratories, all blood samples were tested using established automated systems: BactAlert in Pakistan and BD BACTEC 9050 FX 200 SERIAL NO. FT9356 in Kenya. When a positive signal was identified by the automated systems, samples were subcultures in MacConkey agar and blood agar plates. Non-lactose fermenting colonies on MacConkey agar were biochemically detected for *S*. Typhi and confirmed by *Salmonella* specific anti-sera. For the purposes of this study, patients were classified as typhoid or non-typhoid cases according to the blood culture result. Serum samples from other isolates including *S*.Paratyphi A were not included in the study.

### Sample size and statistical analysis

Based on the reported prevalence of *S*. Typhi in the endemic regions (7% in Pakistan; 5% in Kenya [based on hospital data]) and assuming a low sensitivity and specificity for the RDTs (50% for each), it was estimated that a sample size of 2,900 patients in Pakistan and 2,000 patients in Kenya would be required. This would ensure 200 positive cases and 200 negative cases in Pakistan and 100 positive cases and 100 negative cases in Kenya, enabling the estimation of RDT sensitivity with 80% power to detect a 95% confidence interval (CI) of ±10% in Pakistan and ±14% in Kenya (Wilson method). The retrospective analysis of samples was planned to ensure these targets were met in the event of recruitment difficulties.

The main analysis population was the modified intent-to-test (mITT) population, defined as all patients who were enrolled in the study (intent-to-test [ITT] population) and for whom blood culture test results and at least one RDT test result was available.

Patient characteristics are presented descriptively. RDT sensitivity and specificity values were defined using true positive (TP), true negative (TN), false positive (FP) and false negative (FN) rates, which were calculated using *S*. Typhi blood culture results as the reference standard. Sensitivity was defined as TP / (TP + FN) and specificity defined as TN / (FP + TN). Point estimates for sensitivity and specificity with 95% CIs were calculated using the Wilson method.

Because blood culture has modest sensitivity (estimated at around 60%) to diagnose tyhoid, we perform Bayesian latent class modeling to estimate sensitivity and specificity of the RDTs. This approach leverages prior information about the accuracy of blood culture, along with the joint results of all the RDTs, for estimating each RDT’s accuracy. For tests that had both IgG and IgM results, we selected the better performing isotype from the crude analysis for the latent class analysis. Following previously published methods (12), we defined sensitivity and specificity of each diagnostic by a Beta distribution and used a Gibbs sampler to jointly estimate the sensitivity and specificity, as well as the prevalence of typhoid. We used 100,000 Monte Carlo iterations, discarded the first 50,000, and thinned by 100, reporting median posterior estimates with 95% credible intervals (CrI). We used Beta distributions with α and β equal to 1 for RDT sensitivity, specificity and typhoid prevalence, α and β equal to 57 and 38 for blood culture sensitivity (60% with 95% CI 50–70%), and presumed specificity of >99.99%. Latent class analyses were performed using R.

### Data availability

The authors confirm that data supporting the findings of this study are available within the article.

## Results

### Study population and patient characteristics

Patient enrolment and selection is presented in **Figure 1**. Overall, 3091 patients were enrolled in the study (2980 prospectively and 111 retrospectively), out of which 38 were withdrawn who refused to blood collection after giving consent. 2942 blood culture samples were collected prospectively and 111 samples retrospectively. Of these, 268 were positive for *S*. Typhi (211 from prospective samples and 57 from retrospective samples). Of 2942 samples collected prospectively, 255 were contaminant. Other isolates were *S. Paratyphi A, Escherichia coli, Staphylococcus aureus*, coagulase negative *Staphylococcus, Pseudomonas* spp, *Acinetobacter* spp.

**FIG 1.**
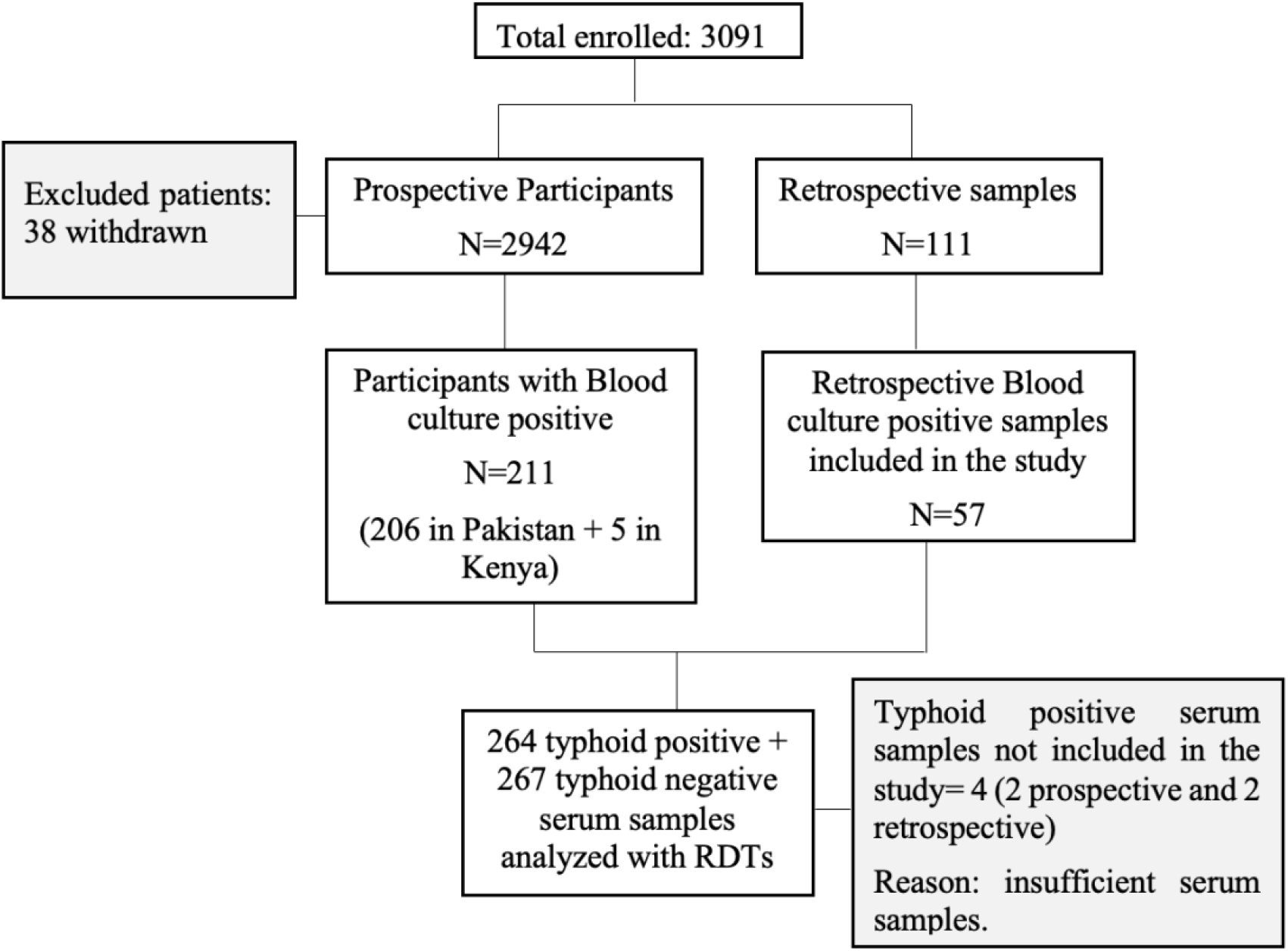
Patient disposition RDT, *S*. Typhi rapid diagnostic test.

Patient characteristics (ITT population) are summarized in **Table 1**. Although most patients were >17 years of age (58%), the large majority of positive *S*. Typhi cases (71.1%) were identified in children <12 years. The proportion of patients who had received a typhoid vaccination was low (4.8% overall), despite an ongoing vaccination campaign in Pakistan.

**TABLE 1.** Patient characteristics, including *S*. Typhi prevalence (ITT population)

|  | **Total** |  | **Typhoid Positive** |  | **Typhoid Negative** |  |
| --- | --- | --- | --- | --- | --- | --- |
|  | n | % | n | % | n | % |
| **Total blood culture** | 2687 | 100 | 211 | 7.9 | 2476 | 92.1 |
| Pakistan | 2155 | 100 | 206 | 9.6 | 1949 | 90.4 |
| Kenya | 532 | 100 | 5 | 0.9 | 527 | 99.1 |
| **Age (mean=22.7)** | 2687 |  | 211 |  | 2476 |  |
| <12 years | 900 | 33.5 | 150 | 71.1 | 750 | 30.3 |
| 12–17 years | 229 | 8.5 | 10 | 4.7 | 219 | 8.8 |
| >17 years | 1558 | 58 | 51 | 24.2 | 1507 | 60.9 |
| **Single-dose TCV**<br>**received\*** | 144 | 4.8 | 15 | 7.1 | 114 | 4.6 |

\*A typhoid vaccination campaign was ongoing in Pakistan during the study.
ITT, intent-to-test; TCV, typhoid conjugate vaccine.

### RDT sensitivity and specificity

In total, 531 serum samples were evaluated using the nine RDTs (264 in positive cases and 267 in negative cases). The sensitivity and specificity of each RDT is summarized in **Figure 2**. Sensitivity values varied widely between the different tests, from 0% (Diaquick Ag cassette) to 78.8% (IgG component of the Typhoid IgG/IgM Combo Rapid Test CE [CTK]). However, IgG tests show the presence of a past infection (13), and the sensitivity of the IgM component of the CTK test (for present infection) was very low (1.5%). The second highest sensitivity (72.7%) was observed with the Enterocheck WB IgM test (Tulip diagnostics). Specificity values showed less variation between RDTs, ranging from 59.2% (IgG component of the Typhoid IgG/IgM Combo Rapid Test CE [CTK]) to 100% (Diaquick Ag cassette, IgM components of the SD Bioline *S*. Typhi IgG/IgM Fast and Typhoid IgG/IgM Combo Rapid Test CE [CTK]).

**FIG 2.**
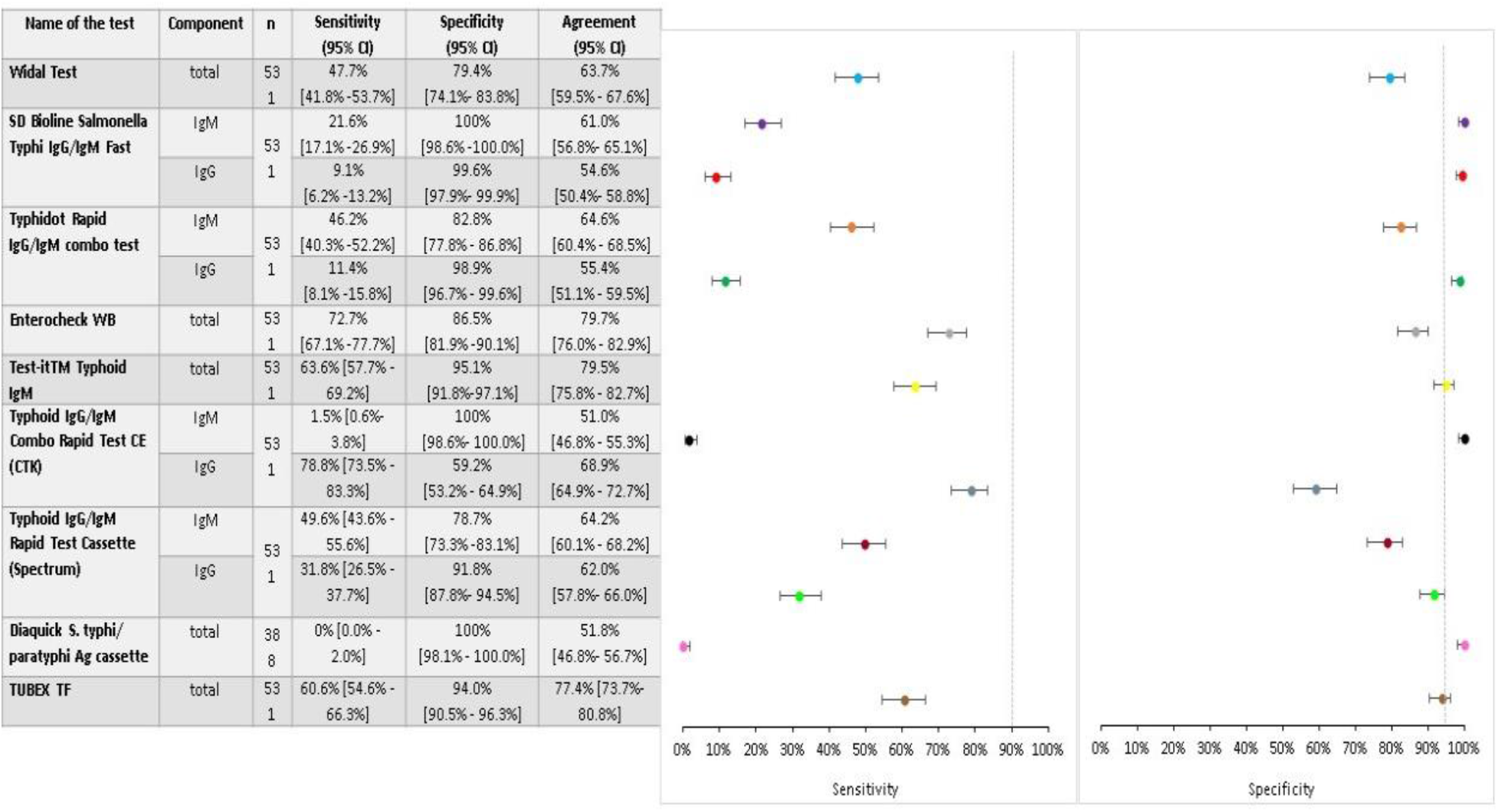
Sensitivity and specificity of nine *S*. Typhi RDTs^*^ (mITT population) *Assessed using blood culture testing to confirm diagnosis. CI, confidence interval (Wilson method); IgG, immunoglobulin G; IgM, immunoglobulin M; mITT, modified intent-to-test; RDT, rapid diagnostic test.

**FIG 3.**
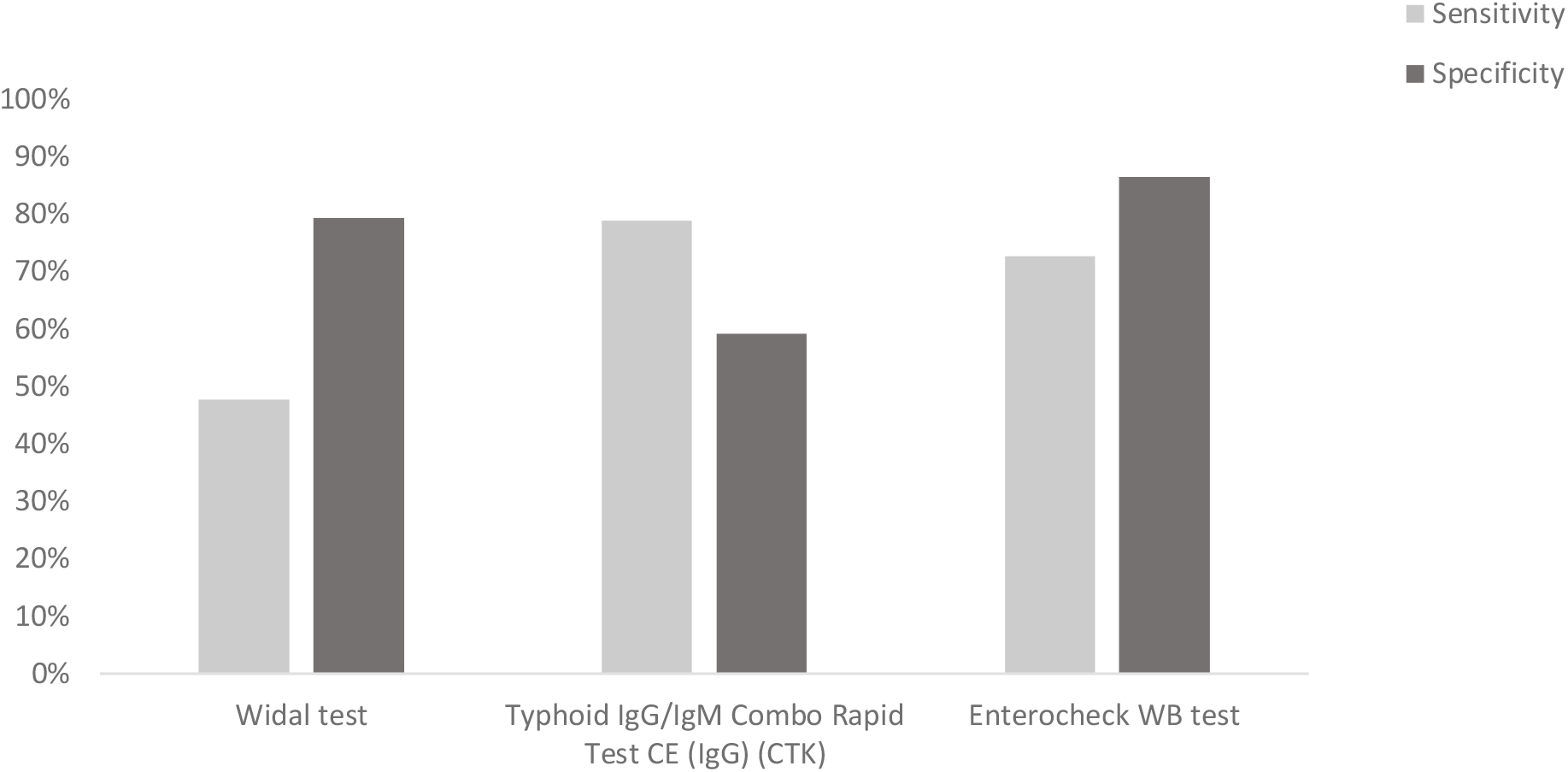
Comparison of Widal test with two best performing RDTs IgG, immunoglobulin G; IgM, immunoglobulin M; RDT, *S*. Typhi rapid diagnostic test.

Overall, the sensitivity and specificity of RDTs was broadly similar (differences <20%) across both endemic regions with three exceptions (**Table 2**): the Typhoid IgG/IgM Rapid Test Cassette showed a greater sensitivity in Pakistan (IgG component: 58.0% *vs* 20.3%; IgM component: 37.6% *vs* 11.9%]), the Test-it™ Typhoid IgM test and Typhidot Rapid IgG/IgM test (IgM component)showed a greater sensitivity in Kenya (81.4% *vs* 58.5% and 93.2% *vs* 32.7% respectively; **Table 2**). Similar to the overall findings, the highest sensitivities were provided by the IgG component of the Typhoid IgG/IgM Combo Rapid Test CE (82.4% in Pakistan) and the Enterocheck WB IgM test (83.1% in Kenya).

**TABLE 2.** Sensitivity and Specificity of *S*. Typhi RDTs stratified by endemic region (Pakistan or Kenya; mITT population)

|  |  | **Pakistan (n=410)** |  |  | **Kenya (n=121)** |  |  |
| --- | --- | --- | --- | --- | --- | --- | --- |
| **Test** | **Component** | **n** | **Sensitivity**<br>**(95% CI)** | **Specificity**<br>**(95% CI)** | **n** | **Sensitivity**<br>**(95% CI)** | **Specificity**<br>**(95% CI)** |
| Widal Test | total | 410 | 48.3%<br>(41.5–55.1) | 75.1%<br>(68.8–80.5) | 121 | 45.8%<br>(33.7–58.3) | 93.5%<br>(84.6–97.5) |
| SD Bioline *S.*<br>*Typhi* IgG/IgM<br>Fast | IgM | 410 | 19.5%<br>(14.7–25.5) | 100%<br>(98.2–100) | 121 | 28.8%<br>(18.8–41.4) | 100%<br>(94.2–100) |
|  | IgG |  | 5.4%<br>(3.0–9.4) | 99.5%<br>(97.3–99.9) |  | 22.0%<br>(13.4–34.1) | 100%<br>(94.2–100) |
| Typhidot Rapid<br>IgG/IgM combo<br>test | IgM | 410 | 32.7%<br>(26.6–39.4) | 93.2%<br>(88.9–95.9) | 121 | 93.2%<br>(83.8–97.3) | 48.4%<br>(36.4–60.6) |
|  | IgG |  | 10.7%<br>(7.2–15.7) | 98.5%<br>(95.8–99.5) |  | 13.6%<br>(7.0–24.5) | 100%<br>(94.2–100) |
| Enterocheck<br>WB | total | 410 | 69.8%<br>(63.2–75.6) | 88.3%<br>(83.2–92.0) | 121 | 83.1%<br>(71.5–90.5) | 80.6%<br>(69.1–88.6) |
| Test-it™<br>Typhoid IgM | total | 410 | 58.5%<br>(51.7–65.1) | 94.6%<br>(90.6–97.0) | 121 | 81.4%<br>(69.6–89.3) | 96.8%<br>(89.0–99.1) |
| Typhoid<br>IgG/IgM Combo | IgM | 410 | 0%<br>(0.0–1.8) | 100%<br>(98.2–100) | 121 | 6.8%<br>(2.7–16.2) | 100%<br>(94.2–100) |
| Rapid Test CE<br>(CTK) | IgG |  | 82.4%<br>(76.6–87.0) | 58.0%<br>(51.2–64.6) |  | 66.1%<br>(53.4–76.9) | 62.9%<br>(50.5–73.8) |
| Typhoid<br>IgG/IgM Rapid<br>Test Cassette<br>(Spectrum) | IgM | 410 | 58.0%<br>(51.2–64.6) | 74.6%<br>(68.3–80.1) | 121 | 20.3%<br>(12.0–32.3) | 91.9%<br>(82.5–96.5) |
|  | IgG |  | 37.6%<br>(31.2–44.4) | 89.8%<br>(84.8–93.2) |  | 11.9%<br>(5.9–22.5) | 98.4%<br>(91.4–99.7) |
| Diaquick S.<br>typhi/paratyphi<br>Ag cassette | total | 387 | 0%<br>(0.0–2.0) | 100%<br>(98.1–100) | 121 | * | * |
| TUBEX TF | total | 410 | 63.4%<br>(56.6–69.7) | 92.2%<br>(87.7–95.1) | 121 | 50.8%<br>(38.4–63.2) | 100%<br>(94.2–100) |

\*\*The Diaquick S. typhi/paratyphi Ag cassette test was not performed in Kenya.
CI, confidence interval; RDT, rapid diagnostic test; IgG, immunoglobulin G; IgM,
immunoglobulin M; mITT, modified intent-to-test.

In latent class modeling, CTK IgG was found to have the highest sensitivity (79.1%) followed by Enterocheck (73.8%) (**Table 3**). Specificity ranged between 63.8% (CTK IgG) and 99.7% (TestIt). The highest overall accuracy was observed with Enterocheck, with 73.8% (95% CrI 68.4–78.9%) sensitivity and 94.5% (95% CrI 90.8–97.2%) specificity.

**TABLE 3.** Latent Class modelling of the test results

| S. n. | Test | Sensitivity |  |  | Specificity |  |  |
| --- | --- | --- | --- | --- | --- | --- | --- |
|  |  | Median | 95% CI |  | Median | 95% CI |  |
| 1 | Blood Culture | 0.831 | 0.789 | 0.867 | 1 | 1 | 1 |
| 2 | CTK IgG | 0.791 | 0.739 | 0.83 | 0.638 | 0.582 | 0.701 |
| 3 | Enterocheck | 0.738 | 0.684 | 0.789 | 0.945 | 0.908 | 0.972 |
| 4 | SD IgM | 0.199 | 0.155 | 0.249 | 0.997 | 0.986 | 1 |
| 5 | Spectrum IgM | 0.497 | 0.44 | 0.552 | 0.821 | 0.77 | 0.863 |
| 6 | TestIt | 0.62 | 0.56 | 0.675 | 0.997 | 0.986 | 1 |
| 7 | Tubex | 0.582 | 0.529 | 0.639 | 0.971 | 0.943 | 0.989 |
| 8 | Typhidot IgM | 0.457 | 0.397 | 0.523 | 0.852 | 0.798 | 0.897 |
| 9 | Widal | 0.486 | 0.425 | 0.545 | 0.837 | 0.784 | 0.881 |
| 10 | Prevalence | 0.548 | 0.504 | 0.591 |  |  |  |

### Secondary outcomes

A biorepository of 389 characterized serum samples (and basic clinical data) was established at the Aga Khan University central laboratory (187 typhoid-positive and 202 typhoid-negative), for use in the development of new tests and the evaluation of emerging technologies in the future via the FIND biobank (https://www.finddx.org/biobank-services/specimen-bank/mrf/). Invalid tests were reported for two of the RDTs, the Test-it™ Typhoid IgM test (0.9% invalid test rate) and the Diaquick Ag cassette (23.8% invalid test rate).

## Discussion

This study evaluated the performance of nine commercially available *S*. Typhi RDTs in two geographically distant endemic regions using a single, established test as a reference standard (blood culture). In general, the RDTs showed poor sensitivity (0–79%) compared with the rigorous standards set by WHO for an effective laboratory test for the diagnosis of typhoid (14). Specificity of the RDTs was higher, but still not always >90%. In particular, the Widal test showed very low sensitivity and specificity (47.7% and 79.4%, respectively) and the SD Bioline *S*. Typhi IgG/IgM Fast test showed very low sensitivity (21.6% for IgM, 9.1% for IgG), but nearly 100% specificity. For the majority of tests the difference between regions was minimal, with only the IgM component of Typhoid IgG/IgM Rapid Test Cassette (spectrum), the Test-it™ Typhoid IgM test and IgM component of Typhidot Rapid IgG/ IgM showing >20% differences in sensitivity (one greater in Pakistan, later two greater in Kenya). Overall, the best performing RDT was the Enterocheck WB, with 72.7% sensitivity and 86.5% specificity. While the Typhoid IgG/IgM Combo Rapid Test CE showed a higher sensitivity for IgG, this typically reflects a previous infection (13), and the sensitivity for acute infection (IgM) was very low.

In general, the findings of this study are supported by those of previous studies (15–23). A study in Bangladesh reported similar findings for the SD Bioline *S*. Typhi IgG/IgM Fast, the Typhoid IgG/IgM Combo Rapid Test CE (IgM component) and Test-it™ Typhoid IgM (15). Specificity findings for the Typhidot Rapid IgG/IgM combo test are also consistent with a recent study conducted in Fiji, as are the Test-it™ Typhoid IgM findings (16). Sensitivity and/or specificity results for the TUBEX TF test were consistent with those reported in studies conducted in various regions, including Bangladesh, Southern Vietnam and Papua New Guenea as well as a systematic review and meta-analysis of studies in several typhoid-endemic countries (9, 17, 18, 20). Finally, the poor sensitivity and specificity showed by the Widal test in this study is in line with established consensus and the findings from many previous studies (6, 8, 9, 21–23).

However, there are also studies for many of the RDTs that show slightly-to-widely contrasting sensitivity and specificity values (15–17, 24–27) probably due to less sample sizes and reference standard differences. Nevertheless, considering the head-to-head nature of this study and the consistent methodology/reference standards employed, together with the broad consensus of the findings across the literature, it is hoped the results presented here may help WHO and health provider decision-making regarding the utility of commercial RDTs in different settings and use cases while clearly highlighting which tests should not be used in practice. The findings further highlight the critical investment in the development of next generation RDTs that could be used at point of care.

Limitations to this study include the use of blood culture as a reference standard, which itself is an imperfect test with limited sensitivity (6). Alternatives to blood culture testing also have their challenges, with bone marrow testing being a complicated and invasive procedure and polymerase chain reaction testing also having limited sensitivity (6). Currently, there is no simple, well-performing, accessible test to use as a gold-standard reference standard. To address this, we used Bayesian latent class modeling to jointly estimate the accuracy of RDTs, using their mutual information and accounting for the imperfect sensitivity of blood culture to better estimate true diagnostic accuracy. This approach has been used in several previous typhoid diagnostics studies (28–30). Another limitation is that RDTs were performed by laboratory staff who had been specifically trained in the appropriate procedures. In routine clinical practice, users would likely follow the manufacturer’s instructions without specialized training, which could potentially mean that differences in RDT ease of use could result in differences in the test results (e.g., invalid test rates). Future real-world evidence studies may provide further insight. Finally, the limited number of retrospective samples in the study precluded comparisons between prospective and retrospective samples to detect any differences (e.g., as a result of Ig deterioration in stored samples). Use of frozen retrospective samples in one of the study sites might have impacted the performance of RDTs. However it is unlikely as apart from IgM component of Typhidot Rapid IgG/IgM, all other test results were comparable between sites. Retesting of the samples for incomparable results were not possible due to lack of samples in retrospective study.

In conclusion, all the *S*. Typhi RDTs evaluated in this study had sensitivity and specificity values that fell below the required levels to be recommended for an accurate diagnosis. However, the Enterocheck WB and Test-it™ Typhoid IgM tests showed the best performances, considerably better than the widely used Widal test, despite not being commonly used in clinical practice. There were minimal differences in RDT performances between endemic regions, with only the Typhoid IgG/IgM Rapid Test Cassette and the Test-it™ Typhoid IgM test showing >20% differences in sensitivity.

Overall, the performance of the *S*. Typhi RDTs in this study highlights the clear need for new developments in the typhoid test landscape, and it is hoped the biorepository of characterized serum samples established at the Aga Khan University central laboratory will help in the development and evaluation of this next generation of tests.

## Data Availability

All data produced in the present work are contained in the manuscript

## Acknowledgments

Editorial assistance was provided by Stuart Wakelin, PhD, supported by FIND – the global alliance for diagnostics. We acknowledge the Director General, KEMRI where the samples from the sites in Kenya were analyzed. We would like to express our gratitude to all fiend officers, laboratory staffs all the patients and other participants of the study. This study was funded by the UK government with a grant to FIND – the global alliance for diagnostics.

